# Variation of malaria dynamics and its relationship to climate in western Kenya during 2008-2019: a wavelet approach

**DOI:** 10.1101/2024.10.31.24316488

**Authors:** Alexis Martin-Makowka, Bryan O. Nyawanda, Anton Beloconi, Godfrey Bigogo, Sammy Khagayi, Stephen Munga, Patrick K. Munywoki, Ina Danquah, Jürg Utzinger, Penelope Vounatsou

**Author notes:** **Corresponding author** Penelope Vounatsou *Swiss Tropical and Public Health Institute, Allschwil, Switzerland and University of Basel, Basel, Switzerland.

## Abstract

Malaria is a vector-borne disease, subject to climate change. The true impact of climate change on malaria dynamics is, however, still debated. Between 2008-2019, we studied patterns of malaria dynamics in a lowland area of western Kenya. We used wavelet analysis to assess the seasonality of monthly malaria incidence and related climatic factors, including air temperature, land surface temperature, rainfall and Niño 3.4 sea surface temperature. We performed a maximal overlap discrete wavelet transform to decompose incidence and climatic factors and fitted bivariate linear regressions to analyse their relationships across time scales. We observed a strong semestrial seasonality of malaria with the emergence of an annual cycle with variation strongly associated with rainfall dynamics. Rainfall emerged as a significant short-term predictor, while temperature contributed more at higher time scales. We found a recent increase in the time lag between climatic factors and their related effects on malaria incidence. This augmentation is related to bed net coverage and El Niño events. Our study underlines the importance of considering long-term time scales when assessing malaria dynamics. The presented wavelet approach could be applicable to other infectious diseases.

## Introduction

Malaria is one of the most deadly infectious diseases and it continues to cause huge public health burden. Understanding its transmission dynamics is crucial for developing effective control and prevention strategies. Recently, a change of transmission dynamics has been observed, and the world incidence has reached a plateau (WHO, 2023). This change in transmission dynamics has been attributed to many factors, including mosquitoes becoming resistant to insecticides, Plasmodium parasites that have become resistant to drugs and the potential influence of climate change (Field and Barros, 2014; Tanser et al., 2003; WHO, 2023).

Malaria is a climate-driven infectious disease. Suitable weather conditions can favour its vector, the *Anopheles* mosquito. Research investigating the impact of climate on malaria incidence showed that key climatic factors are rainfall and temperature (Mordecai et al., 2019; Thomson et al., 2017). The seasonal cycles of malaria incidence closely follow seasonal weather variations. Seasonality refers to the periodicity within a year, that is the patterns and fluctuations in malaria incidence or climatic factors that recur under the 1-year time scale, while multiannual cycles refer to periodicities of length greater than a year. We note that two (or more) periodicities can co-occur, for instance in the presence of a semestrial peak, but every other peak being higher in intensity (presence of a 6-month period together with a 12-month period). Reiner et al. (2015) put forth a systematic review assessing the seasonality of different malaria metrics, and their relationships with climate data. In addition to showing that the best predictors for malaria outbreaks are usually temperature and rainfall, the authors observed that the time lag between a climatic event and an increase in malaria occurrence is highly dependent on the metric, and the area under observation. For instance, the authors found that the time lag between temperature and malaria incidence can vary from 0 to 9 months, while the time lag between rainfall and malaria incidence ranges between 0 and 6 months. Additionally, it was concluded that in most publications, the seasonality of malaria is directly linked to the seasonality of the main climatic factors (usually temperature and rainfall). This conclusion is still valid - most of the publications that considered seasonality investigated the relationship between malaria and climatic factors. However, Pascual et al. (2008) have observed multiannual cycles, inherent to the disease itself and which do not depend on external factors, such as climate. More recently, such cycles have been attributed to global climate drivers such as the El Niño Southern Oscillation (Cazelles et al., 2023).

The aim of this paper was to strengthen the evidence-base about the periodicity of malaria incidence and its dependence/independence on climatic factors, based on a dataset of reported malaria incidence from a western lowland area in Kenya, along with its climate reports.

## Methods and Materials

### Dataset

Malaria cases were extracted from the population-based infectious disease surveillance (PBIDS) system, set up in 2005 by the Kenya Medical Research Institute (KEMRI) and the United States Centers for Disease Control and Prevention (US CDC). The PBIDS is conducted in 33 villages, and follows about 30,000 people residing within a 5km radius from the St. Elizabeth Lwak Mission Hospital in Asembo, Rarieda sub-county, Siaya county, Kenya. Patients with febrile illness symptoms were consented to provide finger prick blood for microscopy. Malaria was diagnosed when *Plasmodium* parasites were detected. We considered malaria cases among children under 5 years of age. Indeed, in our previous analyses using Bayesian methods we observed clearer seasonal patterns compared to older individuals (Nyawanda et al., 2023). Monthly malaria cases among these children from 2008 to 2019 were divided by the person-time (in years) of exposure to obtain the monthly time series of incidence (cases per person-year of observation). More detailed explanations of the dataset, along with a map of the study area, can be found in Nyawanda et al. (2023). Bed net coverage was also reported in the PBIDS. Households were routinely visited for interviews, and all household members were asked whether they slept under a bed net the preceding night. The proportion of household members who used bed nets was computed and aggregated at a monthly time scale.

Rainfall data were obtained via climate hazards group infrared precipitation with station data (CHIRPS). Those data are precise at 5.6 × 5.6 km^2^ spatial and 5 days temporal resolution (Funk et al., 2015). Temperature data were extracted from two different sources. Air temperature was accessed through the ERA5-Land dataset (Muñoz Sabater, 2019), where daily air temperature is measured 2m above ground surface, at a 9 × 9 km^2^ spatial and hourly time resolution. Land surface temperature at day (LSTD) and at night (LSTN) were obtained from the moderate resolution imaging spectroradiometer (MODIS) on board NASA’s Terra and Aqua satellites. Spatial resolution is 1 × 1 km^2^ and temporal resolution is 8 days (Wan et al., 2015). Rainfall, air temperature, LSTD and LSTN were initially computed at their original spatial resolution and averaged per month. Then, the climate data were averaged at the village spatial resolution, and again averaged over all villages, to obtain one value for the whole study area. Missing LSTN values were estimated by averaging the value from the preceding and succeeding months. Land surface temperature (LST) was computed after aggregation as the average of LSTD and LSTN. Both air temperature and LST were selected in the analysis, because they have proven to have different effect on malaria at different time scales. The Niño 3.4 sea surface temperature (SST) index time series was used as an indicator for the El Niño Southern Oscillations (ENSO). ENSO is an important climatic phenomenon known for having an important impact worldwide, and can influence malaria outbreaks (Anyamba et al., 2019; Bouma et al., 1997). Values of the Niño 3.4 SST index are monthly averaged SSTs of the Niño 3.4 oceanic area, which is located between longitudes 120-170W and latitudes 5S-5N. The Niño 3.4 SST data were extracted from the HadISST1 dataset (Rayner et al., 2003).

### Analysis of periodicity

Seasonalities of malaria incidence and climate time series were analysed using wavelets, specifically through a continuous wavelet transform. The overall concept of wavelets is related to the search of frequencies (or inversely, periodicities/seasonalities) in a time series. They offer powerful tools for the investigation of non-stationary processes, rendering them a suitable approach for studying infectious diseases and climate data (Cazelles et al., 2018).

A wavelet is a function representing a small oscillation localized in time. In the process of the continuous wavelet transform, we used a wavelet of reference (a *mother* wavelet, in our case a Morlet wavelet) that we scaled and shifted. We applied a transform between the time series of interest and the wavelet obtained by scaling and shifting. For each of the scale coefficient *s* and the shift coefficient τ, the transform carried information on how strong the seasonality of period *s* was around time point τ in the time series. The wavelet power spectrum of the transform was the quantity carrying this information, and had been plotted as a heat map. The force of the periodicity was assessed at all scales/periods and all time points within the study duration. Statistical tests were employed to assess the true seasonal dynamics. Random lag-1 autoregressive models were simulated to generate a baseline distribution for the wavelet power spectrum. This process allowed us to test the wavelet power spectrum obtained from our data against the null hypothesis of ‘no seasonal pattern at this time point and period’ (Torrence and Compo, 1998). Taking the average wavelet power spectrum along a certain period gave the average power spectrum, an unbiased variant of the Fourier transform, which could also be tested for periodicity at different period values using simulations. A more complete description of the continuous wavelet transform can be found in Cazelles et al. (2007), and in the description of *WaveletComp*, the R-package used for computation (Rosch and Schmidbauer, 2018).

In a similar way, we obtained information on the common, joint seasonal patterns of two time series: the malaria incidence and each of the climatic factors. For this, we used the cross-wavelet transform. The continuous wavelet transform was applied to both time series and the outputs were merged to compare the seasonal dynamics between the two time series. A wavelet power spectrum was produced and plotted as a heat map. A statistical test to evaluate the true joint seasonality was done similarly to the single time series case. The cross-wavelet power spectrum was averaged into an average power spectrum and tested for significance.

### Phase differences

In addition to assessing the joint seasonal dynamics, the cross-wavelet transform was applied to two time series to determine the phase differences between them. The phase differences carried information on the time between a peak in the first time series (the leading) and the closest following peak of the other time series (the lagging). The phase difference angle that is output from a cross-wavelet transform takes a value of *θ* ∈ [0, 2π). This angle represents the radial distance between two peaks. As an example, an angle of *θ* = 0 means synchronicity (time series are in-phase), and an angle of *θ* = π means that the time series are anti-phase (Fig. 1).

**Figure 1.**
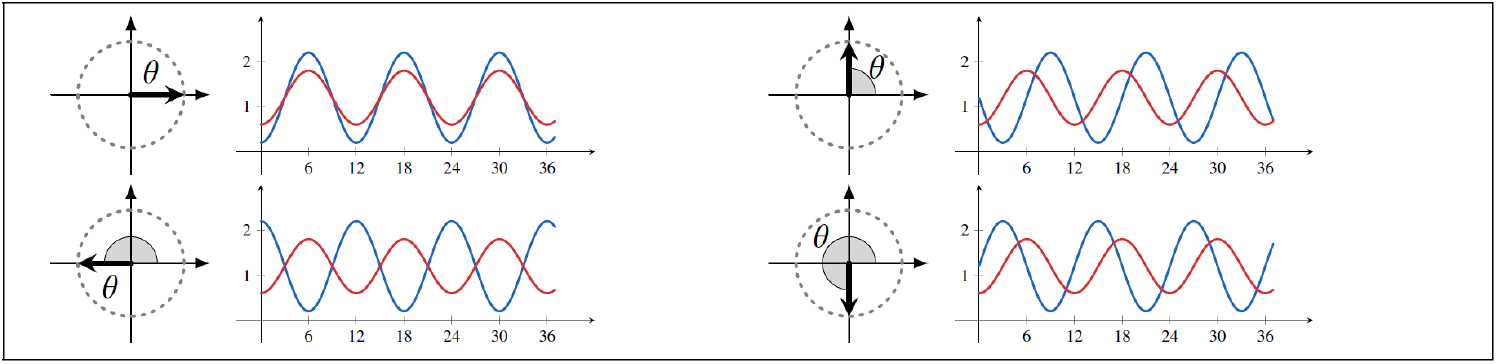
Relationship between the phase difference angles and the time series dynamics. When both time series are in phase, the phase difference angle is *θ* = 0 (upper left). When the red time series is ahead of the blue time series by a quarter of the period (3 units over a period of 12), the value of the phase difference angle is one quarter of the total circle, that is *θ* = π/2 (upper right). When both time series are anti-phase, the phase difference angle is *θ* = *π* (bottom left). When blue time series is behind by three quarter of period (here, 9 over 12 units), *θ* = 3π/2 (bottom right). We observe that we cannot distinguish mathematically with the situation where time series in blue is actually leading by one quarter. In our case, however, malaria incidence always follows the effect of climatic variables and the causality direction is clear. Hence, we avoid negative values when speaking about phase difference in months.

Since the definition of phase difference strongly relies on the notion of periodicity, we could only compute reliable phase differences when both time series displayed a seasonal pattern at this period. Practically, this means that phase differences were only computed for time points and periods where joint periodicity was tested significant; they were plotted directly in the cross-wavelet power spectrum heat map within confidence areas, and took the shape of arrows displaying the angle stated above.

### Time scales decomposition

The direction of the relationship by time scale was obtained using the maximal overlap discrete wavelet transform. Differently from the continuous wavelet transform, the maximal overlap discrete wavelet transform does not compute the periodicity of a time series at all periods. Instead, it parsimoniously decomposes the time series into different subseries, each having a specific periodicity. In other words, it extracts from an original time series its different time scales components, each subseries having its own periodicity. By applying a maximal overlap discrete wavelet transform with a depth level of 3 on a single time series, we obtained four subseries: *D*_1_, *D*_2_, *D*_3_ and *S*_3_. *D*_1_ was the subseries of the lower period, while *D*_3_ was the subseries of the greater period. *S*_3_ retained the trend of the time series. Captured time scales are summarised in Table 1.

The four subseries summed up to the original time series, i.e.

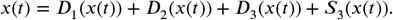

**Table 1.** Time scales obtained after a maximal overlap discrete wavelet transform of depth 3 of an input time series.

|  |  |
| --- | --- |
| $D_1$ | 1- to 4-month periodicities (approximately trimestrial seasonality) |
| $D_2$ | 5- to 8-month periodicities (approximately semestrial seasonality) |
| $D_3$ | 9- to 16-month periodicities (approximately annual seasonality) |
| $S_3$ | 16-month and longer periodicities (trend) |

A more comprehensive description of the method can be found in Appendix 1, and a detailed description of the maximal overlap discrete wavelet transform can be found in Percival and Walden (2000). Our study used the Coiflet wavelet filter of length 30. This wavelet filter had proven efficient over some artificially simulated data of the same nature as the data we studied.

### Multiresolution analysis

After decomposing the time series into multiple subseries, we investigated the bivariate, linear relationships between the subseries of climatic factors and the subseries of malaria incidence. This was done by fitting a linear regression between one climatic subseries and the subseries of malaria incidence at the same time scale. The predictor, that is the climatic subseries was standardized and shifted by a certain lag. The lags were comprised between 0 and a maximal meaningful period at this time scale: 4 for the time scale *D*_1_, 8 for the other time scales.

Decomposing a time series across different time scales generated highly redundant and repetitive subseries. Hence, shifting a climatic subseries by a lag of half the time scale brought the exact opposite results, with linear regression slopes having the opposite sign. To avoid this situation, we selected the lag according to the best fit (lowest Akaike information criterion [AIC]), but when we faced the situation of two different lags generating almost the same fit, we parsimoniously selected the smallest lag.

### Lag variations

The results of the multiresolution analysis were used to turn the phase differences, defined in a previous section, into a dynamic evolution of the lags. We explicitly obtained the phase differences by selecting a slice at a given period, and we computed the phase differences at this period for all time points. Hence, we obtained phase differences function of time. The phase difference angles were converted into months using the formula

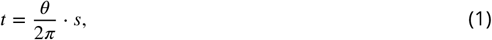

where *t* is the phase difference in months, *θ* is the phase difference as an angle and *s* is the period under investigation. This phase difference in months can be turned into lags if and only if the direction of the relationship between the two variables is known at this period. A positive relationship implies that the lag is equal to the phase difference in months. A negative relationship requires to adjust the phase difference by half a period.

The variations of the lags were investigated using linear regressions between the lag variations and ‘crude’ climatic variables. Computing linear models between all climatic-intervention time series (including bed net coverage) and all lag variations of air temperature, LST and rainfall generated 18 linear regressions. We kept the two models with the lowest AIC.

## Results

### Analysis of periodicity

Inspection of the malaria incidence time series in Figure 2A showed highly seasonal patterns, with repetitive peaks around June and January, which are the months that follow Kenya’s long and short rainy seasons respectively. The incidence declined over time from 2008 to 2015, with 2015 showing the lowest total yearly incidence. Subsequently, a shift was observed in the regime, with higher peaks in June, while peaks around January decreased in size. The wavelet power spectrum plot in Figure 3A tended to confirm this dynamic: malaria incidence showed a clear and statistically significant 6-month periodicity (semestrial seasonality) almost continuously within the whole study duration, with a gap in 2018. There was a slightly stronger 12-month periodicity (annual seasonality) appearing around 2017-2018, but this seasonality lacked significance. When averaged, the only statistically significant seasonality was the 6-month periodicity.

**Figure 2.**
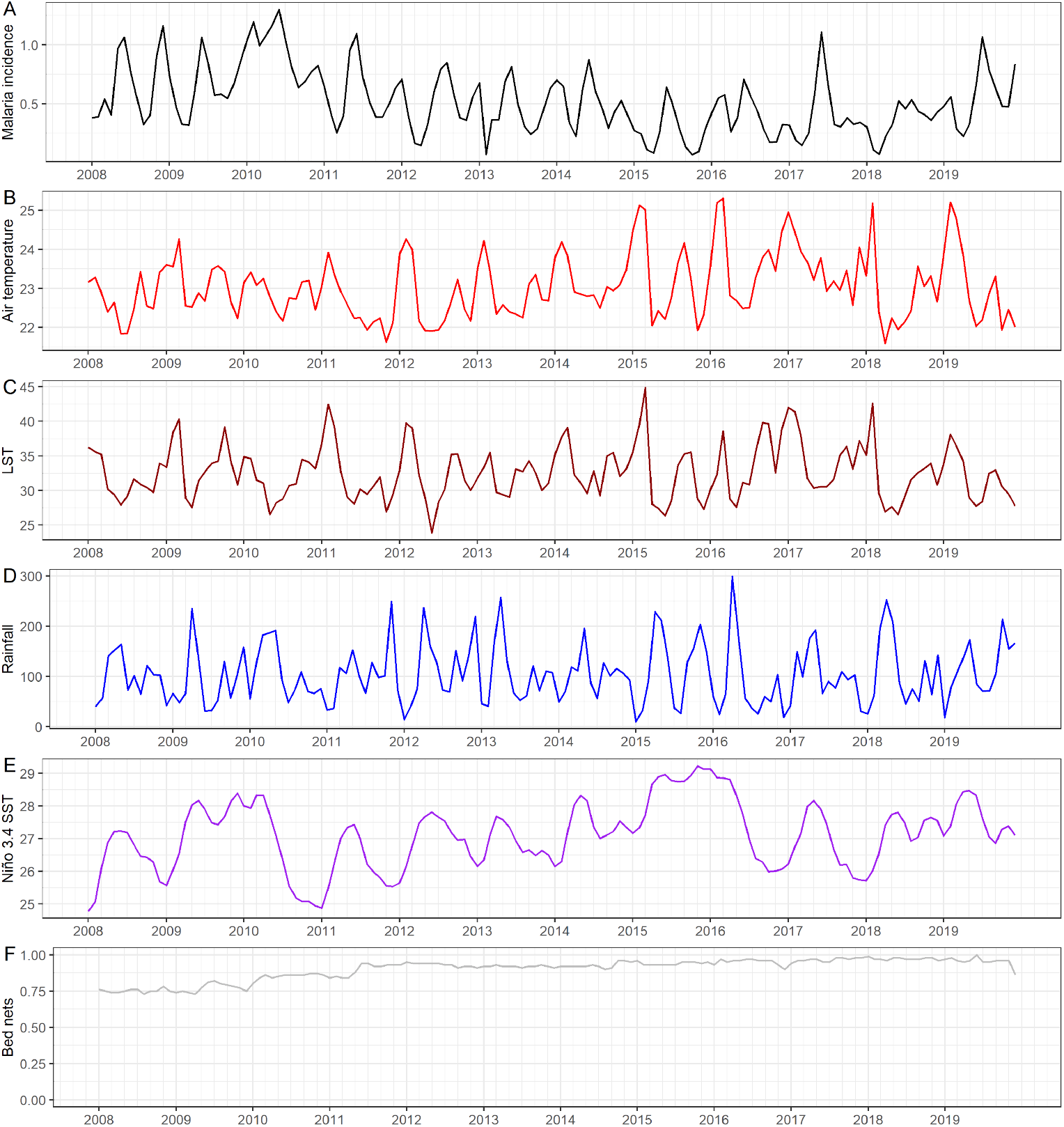
Time series plots (2008-2019) of the different variables of our study. Malaria incidence (A) is computed in terms of cases per person-year, air temperature (B) and LST (C) are in °C, rainfall (D) units are in mm of precipitation, Niño 3.4 SST (E) is a sea surface temperature in °C, and bed net coverage (F) is a percentage of visited households where bed nets were effectively used.

**Figure 3.**
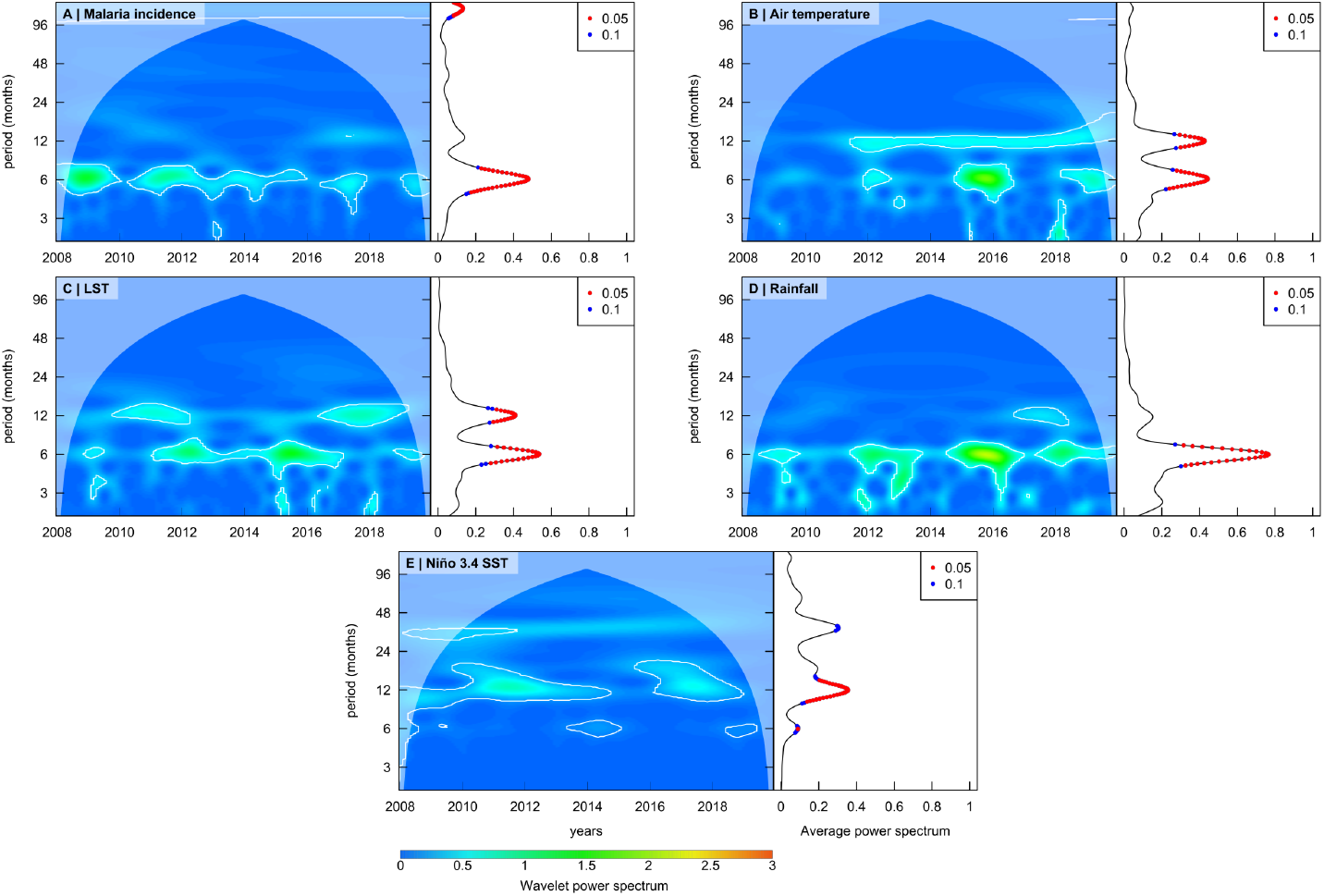
Wavelet power spectra heat maps (left) and average wavelet spectra (right) of malaria incidence (A), air temperature (B), LST (C), rainfall (D) and Niño 3.4 SST (E). Colours represent the spectrum itself, while white contour lines delineate the regions where periodicity is assessed at the 90% confidence threshold, against red noise. Light blue areas on top of the maps represent the cone of influence, where periods are too high and time is too close to time series bounds to be able to assess seasonalities. Solid points in the average power spectrum are periods tested at the 95% (red) or 90% (blue) confidence threshold.

For air temperature, time series plot showed semestrial regular peaks around February and September (Fig. 2B). There was a tendency to switch to one single annual peak in February, from 2015 onwards. The continuous wavelet transform showed that there was no significant pattern of any kind up to 2012 (Fig. 3B). From that year onwards, a strong and uninterrupted 12-month period pattern appeared, along with a clear and more punctual 6-month cycle around the years 2012, 2016 and 2019. Both seasonalities were significant when averaged.

There was a higher variation of seasonality in the case of LST (Fig. 2C). Despite the peaks that regularly occurred in March and October, the continuous wavelet transform revealed highly noncontinuous seasonalities (Fig. 3C). A 6-month period pattern was found in 2009, from 2011 to 2016, and in 2019. A 12-month seasonality occurred from 2009 to 2012 and from 2016 to 2019. Both seasonalities showed statistical significance when averaged.

Rainfall presented high peaks in precipitation in April-May, often followed by two usually lower peaks in September and November (Fig. 2D). The wavelet power spectrum displayed significant seasonalities around the 6-month period, and additionally around the 12-month period in 2016-2018 (Fig. 3D). The 12-month seasonality tested significant when accounting for time, but not when averaged over time. The 6-month period tested significant when averaged.

In contrast with the other climatic variables, Niño 3.4 SST presented higher seasonalities (Fig. 2E). In addition to the regular annual peak around May, confirmed by the wavelet power spectrum throughout the time series but in 2015, we also observed an overall trend (Fig. 3E). This trend is assessed by the wavelet power spectrum as being a significant 36-month seasonality from 2008 up to the end of 2011. The wavelet power spectrum also revealed some semestrial seasonalities around 2014 and 2019. The average power spectrum showed statistically significant 6-month, 12-month and 36-month periods.

Inspection of the bed net coverage time series in Figure 2F was a bit different from the other time series, because there is *a priori* less reason to believe in the presence of any seasonal pattern than for climatic factors. A key observation was, however, the increase in bed net coverage over the study period, increasing from 76% in 2008 to almost 100% in early 2019.

Figure 4 shows the heat maps of the obtained cross-wavelet power spectra. We observed that air temperature along with malaria incidence exhibited a significant and almost uninterrupted joint 6-month seasonality (Fig. 4A). We also observed joint 12-month seasonalities in 2011 and from 2016 onwards. Both periods were tested significant at the 95% confidence threshold in the average power spectrum. Analysis of phase difference (arrows) revealed that a peak in temperature precedes a peak of incidence by around 3-5 months for the 6-month period. They also carried information of a 5 to 6 months delay between these two peaks at the 12-month period.

**Figure 4.**
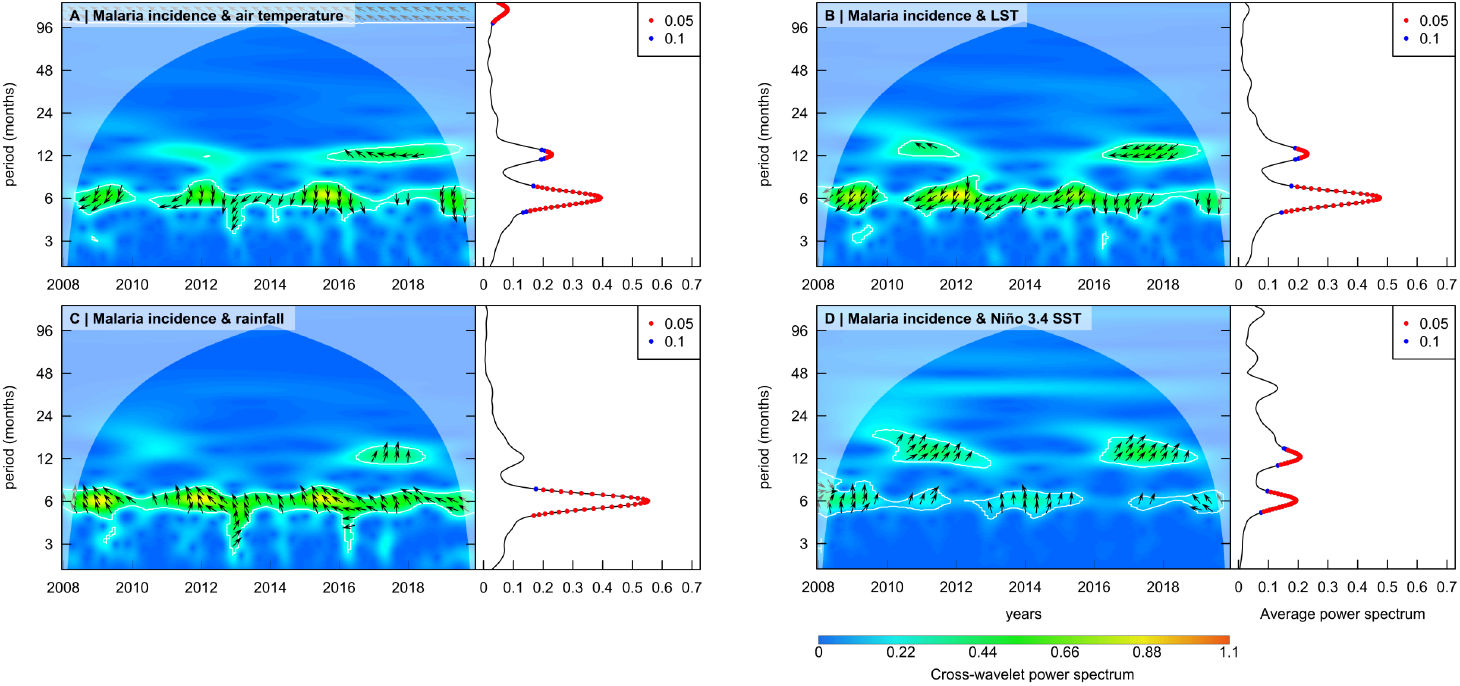
Cross-wavelet power spectra heat map (left) and average power spectra (right) of malaria incidence related to air temperature (A), LST (B), rainfall (C) and Niño 3.4 SST (D). Colours represent the spectrum itself, while white contour lines delineate the regions where periodicity is assessed at the 90% confidence threshold, against two simulated time series following red noise processes. Light blue areas on top of the maps represent the cone of influence, where periods are too high and time is too close to time series bounds to be able to assess seasonalities. Solid points in the average power spectrum are also periods tested at the 95% (red) or 90% (blue) confidence threshold. Arrows represent the phase differences between the climatic time series and the malaria incidence.

The LST cross-wavelet spectrum revealed the same joint dynamics with incidence (Fig. 4B). The 6-month seasonality showed statistical significance during the whole study duration but a small interruption at the beginning of 2010, while the 12-month seasonality was only significant in 2010-2011 and from 2016 onwards. Both periods were tested significant at the 95% confidence threshold in the average power spectrum. The phase differences for the 6-month period oscillated between 3.5 and 4.5 months. The phase differences for the 12-month period were about 4 months between 2010 and 2011, but around 6.5 months from 2016 onwards.

For rainfall, we obtained a clear and continuous joint seasonal pattern with incidence at the 6-month period (Fig. 4C). A common 12-month seasonality was also observed between 2016 and 2018, but this period showed no statistical significance in the average power spectrum. For both time scales, arrows indicated phase differences of approximately 2.5 months.

Niño 3.4 SST showed less clear common seasonality with malaria incidence (Fig. 4D). A common trend was observed at the 6-month period, with gaps in 2012 and 2016. An additional 12-month period joint pattern was observed in 2010-2013 and from 2016 onwards. These two seasonalities showed statistical significance in the average power spectrum. The phase differences were around 1 and 2 at the 6-month period, and around 1.5 at the 12-month period.

### Bivariate relationships across time scales

Figure 5 shows an example of a maximal overlap discrete wavelet transform-decomposed time series (malaria incidence). The decomposition of the climatic time series can be found in the Appendix 2. The full table of results of the multiresolution analysis displaying the results for all lags can be found in Appendix 3. For clarity, we have summarised in Table 2 the results with the selected lags only.

**Table 2.** Results of the bivariate linear regressions between the malaria incidence time series and the predictors, i.e. the climatic time series at different time scales. The climatic subseries obtained through maximal overlap discrete wavelet transform have been standardized before computation of the model. The slope coefficient assesses the direction of the relationship. The numbers in brackets represent the 95% confidence interval bounds for the slope coefficient. The lags for the lowest time scale (*D*_1_, 1-to 4-month period) have been restricted to a maximum of 4, to avoid lags greater than the time scale. The lags for other time scales have been restricted to a maximum of 8.

| Time scale | Covariate | Selected lag | Slope coefficient | 95% confidence interval |
| --- | --- | --- | --- | --- |
| $D_1$ 1-4 mo. | Air temp | 2 | -0.02 | (-0.03, -0.01) |
| $D_1$ 1-4 mo. | LST | 2 | -0.02 | (-0.03, -0.01) |
| $D_1$ 1-4 mo. | Rainfall | No lag with significant fit | | |
| $D_1$ 1-4 mo. | Niño 3.4 SST | 2 | 0.03 | (0.02, 0.04) |
| $D_2$ 5-8 mo. | Air temp | 1 | -0.1 | (-0.12, -0.08) |
| $D_2$ 5-8 mo. | LST | 1 | -0.13 | (-0.14, -0.11) |
| $D_2$ 5-8 mo. | Rainfall | 2 | 0.12 | (0.10, 0.14) |
| $D_2$ 5-8 mo. | Niño 3.4 SST | 2 | 0.11 | (0.09, 0.13) |
| $D_3$ 9-16 mo. | Air temp | 0 | -0.07 | (-0.08, -0.05) |
| $D_3$ 9-16 mo. | LST | 0 | -0.07 | (-0.08, -0.06) |
| $D_3$ 9-16 mo. | Rainfall | 2 | 0.07 | (0.06, 0.08) |
| $D_3$ 9-16 mo. | Niño 3.4 SST | 2 | 0.07 | (0.06, 0.08) |
| $S_3$ 16+ mo. | Air temp | 0 | -0.07 | (-0.09, -0.04) |
| $S_3$ 16+ mo. | LST | 0 | -0.04 | (-0.07, -0.01) |
| $S_3$ 16+ mo. | Rainfall | No lag with significant fit | | |
| $S_3$ 16+ mo. | Niño 3.4 SST | 0 | -0.04 | (-0.07, -0.01) |

**Figure 5.**
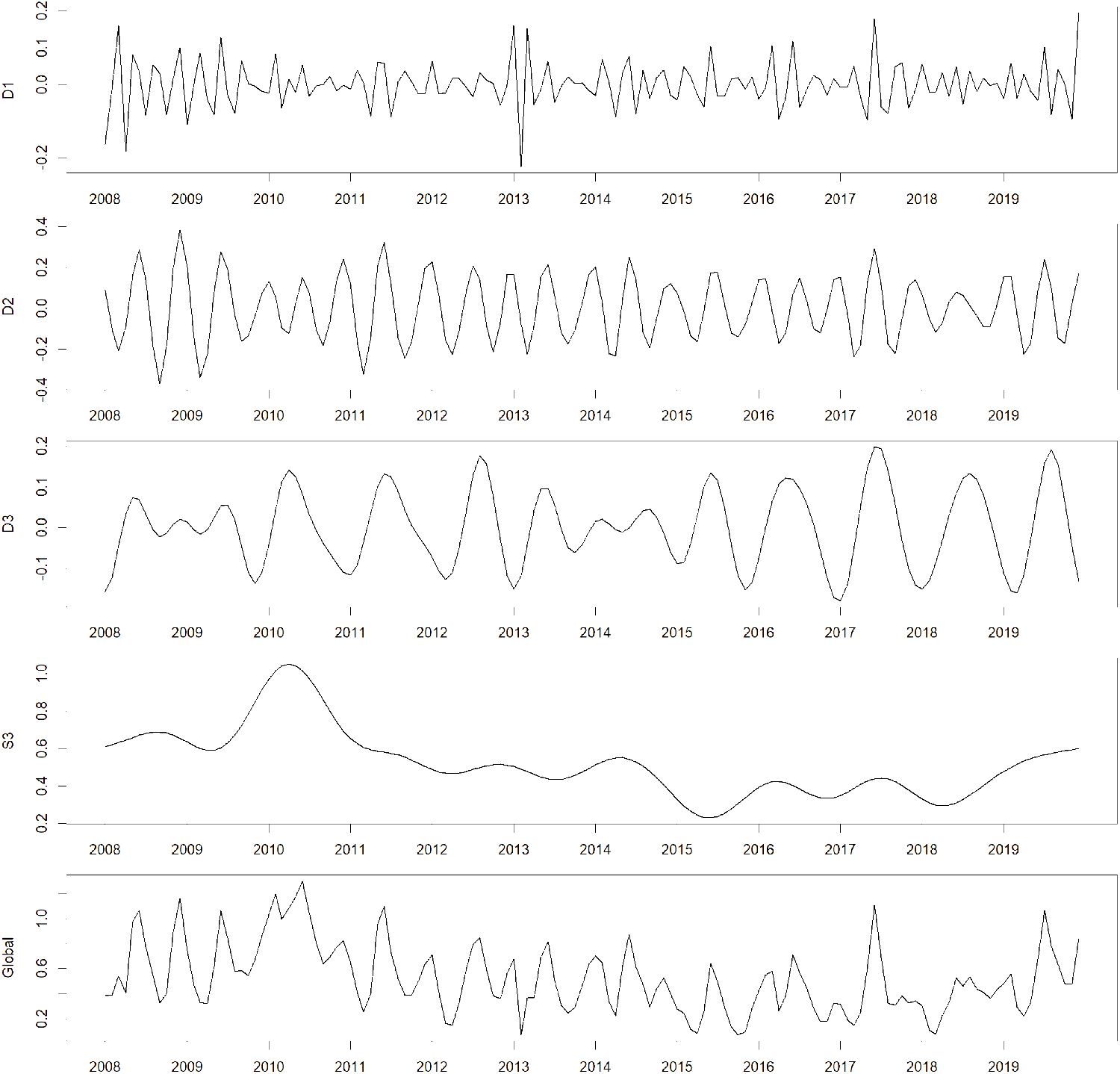
Time series decomposition of the malaria incidence time series using the maximal overlap discrete wavelet transform. The global original time series is plotted at the bottom, while the different subseries are plotted above. The subseries *D*_1_ corresponds to time scales of 1- to 4-month, *D*_2_: 4-to 8-month, *D*_3_: 8-to 16-month and *S*_3_ retains seasonalities of periods above 16-month.

### Lag variations

The phase differences obtained by the cross-wavelet transforms were turned into lags, allowing us to plot the evolution of the lags over time. Phase differences can only be obtained at periods and times where joint periodicity is assessed significant. We extracted the phase differences for the variables air temperature, LST and rainfall at the 6-month period. Rainfall being positively correlated with malaria incidence at the 6-month period, the phase differences (‘peak-to-peak’ elapsing time) were exactly the lag. For both air temperature and LST, they were negatively correlated to malaria incidence at this time scale; we added half-a-period (3 months) to the phase difference to obtain the lag. Figure 6 shows the evolution of the lag of these three variables over the course of the study period.

**Figure 6.**
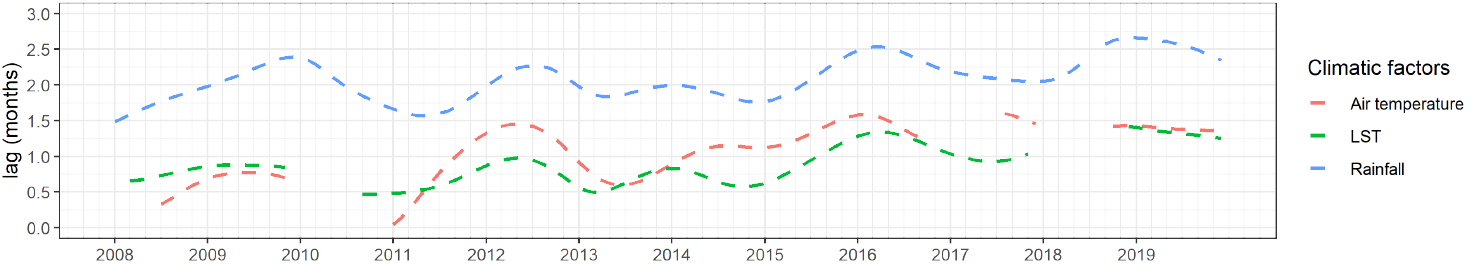
Evolution of the lags obtained through the phase differences. The phase differences of the climatic factors related to malaria incidence have been computed at a period of 6-month. Phase differences have been converted into lags by using the method described in the Methods section.

For the three climatic variables, we observed an upward trend in the lags from 2008 to 2019. Air temperature and LST lags fluctuated between 0.5 and 1.5 months, with interruptions in 2010 and 2018, when the joint periodicity was not significant. The lag between rainfall and malaria incidence varied from 1.5 months to 2.5 months over the course of the study.

The first of these two models was the linear regression of the LST lag variation against the values of the Niño 3.4 SST time series. The regression model showed a positive estimate of 0.11 for the slope coefficient, which was statistically significant. The p-value for both the intercept and the slope coefficient were below 0.1%. The *R*^2^ coefficient was 0.2 and AIC was -0.12 (Table 3). The model with the second lowest AIC was the regression of LST lag against the bed nets occupancy. Models showed a positive estimate of 1.41 for the slope coefficient, which was statistically significant. The p-value for the slope coefficient was below 0.1%, and had a value of 15.7% for the intercept. The *R*^2^ coefficient was 0.15 and AIC was 7.1 (Table 3).

**Table 3.**
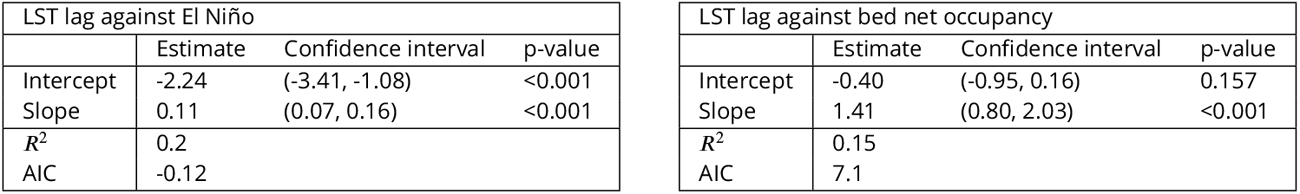
Results of the linear regressions of the lag evolution time series against the climate time series. The time series of the lag evolution of air temperature, LST and rainfall have been fitted with climatic and intervention variables time series: air temperature, LST, rainfall, Niño 3.4 SST and bed net coverage. Only the two models with the least AIC are displayed here.

| LST lag against El Niño |  |  |  | LST lag against bed net occupancy |  |  |  |
| --- | --- | --- | --- | --- | --- | --- | --- |
|  | Estimate | Confidence interval | p-value |  | Estimate | Confidence interval | p-value |
| Intercept | -2.24 | (-3.41, -1.08) | <0.001 | Intercept | -0.40 | (-0.95, 0.16) | 0.157 |
| Slope | 0.11 | (0.07, 0.16) | <0.001 | Slope | 1.41 | (0.80, 2.03) | <0.001 |
| $R^2$ | 0.2 | | | $R^2$ | 0.15 | | |
| AIC | -0.12 |  |  | AIC | 7.1 |  |  |

## Discussion

Climatic factors such as rainfall and temperature are recognized as driving the seasonality of malaria transmission. However, cycles lasting longer than 1 year might be due to either the internal dynamics of malaria or alternative climatic factors; the effect of climatic factors might be altered across time scales. The overall malaria dynamics and the elapsing lag time between the occurrence of a climatic event and its related effect on malaria incidence might change over time. In this study, we developed wavelet-based analyses to assess the seasonal patterns of malaria incidence and its related climatic effects in the lowlands of Siaya county in the western part of Kenya during 2008–2019, using the PBIDS dataset. We decomposed the relationship (force and direction) between the climatic factors and the malaria incidence. We investigated the evolution of the time lag between the climatic factors and the malaria incidence over the course of the study. These analyses enabled us to measure the variations of periodicity over time and the evolution of the climate-malaria incidence lags along to quantify the changes in climate-related effects on malaria across time scales.

In the analysis of periodicity section, we employed the continuous wavelet transform and the cross-wavelet transform to perform these analyses. A key finding of our study is that the malaria incidence in Siaya County exhibited a dominant 6-month seasonal cycle between 2008 and 2019. Towards the end of the study period in 2019, a 12-month seasonality was apparent, although it did not reach statistical significance. This closely aligns with the cycles of rainfall. Our findings are consistent with previous studies highlighting the rain-driven nature of malaria transmission (Boadu, 2019; Reiner et al., 2015). The effect of rainfall on malaria is complex, as rains support the emergence of small water bodies favourable to the aquatic stages of the mosquito’s life cycle. On the other hand, heavy rains are known to flush away mosquito larvae (Lafferty, 2009). Both air temperature and LST exhibited a 12-month (annual) seasonality. This result is not striking as the temperature is very likely to fluctuate annually. However, this pattern was not strongly reflected in malaria incidence, and rainfall seems to be a better predictor of malaria than temperature. The periodicity of Niño 3.4 SST showed considerable variation over time, including high-scale patterns, such as 36-month cycles. Hence, El Niño offers an explanation for the multiyear cycles of malaria reported in previous studies (Pascual et al., 2008).

The exploration of the bivariate relationships between climate and malaria incidence across time scales has required a multi-resolution analysis, using the maximal overlap discrete wavelet transform. We observed discrepancies in the strength of relationships across time scales, as evidenced by the differences in slope coefficients. As predictor, climatic subseries were standardized before the regression, it supports that malaria incidence exhibits a stronger seasonality or trend at specific time scales. Additionally, it also highlights that the relationship between climate and malaria is itself weaker at certain time scales. Notably, the time scale with the strongest relationship for all variables was found to be *D*_2_ (5-to 8-month periods), followed by *D*_3_ (9-to 16-month periods), aligning with the results obtained from continuous and cross-wavelet transforms. It is interesting to note that the trend (*S*_3_, 16-month periods and longer) coefficient are greater than the finest time scale (*D*_1_, 1-to 4-month periods). This suggests that minor climate variations have a limited impact on malaria incidence at shorter time scales, whereas climatic trends may play a more substantial role in shaping long-term malaria dynamics.

The relationship between temperature and malaria incidence consistently displayed a negative correlation across all time scales. Both air temperature and LST exhibited similar time lags, ranging from 0 to 2 months. This direction of the relationship aligns with expectations: in lowlands, higher temperatures are associated with lower malaria transmission due to the climate being too warm for mosquitoes to optimally reproduce (Beloconi et al., 2023; Nyawanda et al., 2023; Wang et al., 2022). It is interesting to note that at shorter time scales (*D*_2_, 5-to 8-month periods), LST had a greater impact than air temperature, while the opposite was observed at the highest time scale (*S*_3_, 16-month periods and longer). However, we did not find any clear explanation for this change in dynamics across these two time scales.

Rainfall showed a positive relationship with a lag of 2 months, except at both the lowest and the highest time scales, where rainfall became statistically non-significant at any lag. The lag of 2 months and the positive relationship are in line with previous analyses conducted on this dataset (Beloconi et al., 2023; Nyawanda et al., 2023) and are supported by general knowledge of the effect of rainfall on malaria transmission (Reiner et al., 2015). The absence of relationship at the small time scales might indicate that small and disparate rains are not sufficient to impact the development of mosquitoes - or, at the opposite, that an intense and sudden rainfall can flush away the larvae. The absence of relationship at higher time scales is most likely due to the absence of a clear trend of rainfall over the years, while malaria incidence time series shows a clear decreasing trend. Interestingly, despite the significant contribution of rainfall to malaria incidence, it is not the factor that drives the changes in malaria dynamics at high time scales, with the latter being more likely to be driven by changes in the trend of temperature. In other words, while a lower share of malaria cases can be attributed to temperature rather than rainfall, it might be the upward trend of temperature that affects the change in malaria dynamic at the high time scales.

The relationship between Niño 3.4 SST and malaria incidence is significant across all time scales. El Niño or la Niña events are defined when the Niño 3.4 SST exceeds ± 0.4°C for a period of 6 months or longer (Trenberth, 1997). During El Niño events, heavy rains are typically observed in Kenya in November and December (Ototo et al., 2011). While El Niño is known to have an impact at inter-annual scales (Bouma et al., 1997), the prediction ability at shorter time scales may be attributed more to coincidence with other climatic variables than to a clear and real impact of SST on malaria incidence in Siaya.

The decomposition of the time series into subseries and the ensuing linear regression were meant to give the direction of the relationship between climatic factors and malaria incidence at different time scales. This has been used to adjust the phase differences, turn them into lags, and study their evolution over time. The analysis of the lag evolution was performed for the air temperature, LST, and rainfall; the only selected period was the 6-month cycle, as the other periods did not exhibit sufficiently significant joint seasonalities between climatic factors and malaria incidence, and joint seasonal patterns are necessary to compute the phase differences. The most striking finding of this analysis is the substantial increase in the time lag between these climatic variables and malaria incidence. The elapsing time between the occurrence of a peak of intensity in one climatic variable and its resulting peak on malaria incidence has more than doubled in certain cases. This highlights a real change in dynamics between climate and malaria and, to our knowledge, has not been reported before. We found a weak relationship between this increase of time lag and the values of the Niño 3.4 SST, as well as with the bed net coverage. However, there is no evidence of causality. Hence, further research is needed before drawing more definitive conclusions.

It is important to recognize a few limitations inherent in our analysis. First, regarding the multiresolution analysis, although it addresses the non-stationary dynamic of the data, our approach was restricted to linear and bivariate analyses. This is a significant limitation given that the effects of variables such as temperature and rainfall on malaria transmission are known to be nonlinear (Mordecai et al., 2019). Secondly, both temperature and rainfall are strong predictors of malaria transmission risk (Kapwata et al., 2021). However, performing a robust multivariate analysis using this methodology is challenging due to the particularly strong periodicity of the subseries.

The outcomes of our analysis offer several implications for public health practice. It is crucial to consider long-term timescales when planning malaria control strategies, as the transmission dynamics of malaria are continually evolving. Public health initiatives should therefore plan for anticipated changes over time. Monitoring climate variability remains a highly effective approach for anticipating malaria outbreaks and developing early-warning systems, which enable more proactive and timely response with malaria prevention and control efforts.

Taken together, our findings indicate that seasonality in malaria incidence is strongly influenced by seasonality in rainfall, while changes in dynamic patterns are more likely driven by other climatic factors that exhibit clearer trends, such as temperature and Niño 3.4 SST. Moreover, our study revealed an increase in the lag duration of some climatic variables, which was associated with bed net coverage and Niño 3.4 SST, although no evidence of causality was found. The relevance of our findings extends to the development of early warning systems and climate change adaptation strategies. Our study underlines the importance of considering long-term time scales when assessing malaria dynamics and highlights the complex interplay between climate and malaria. By improving our understanding of these relationships, we can enhance preparedness and implement targeted interventions to mitigate the impact of malaria on vulnerable populations.

## Ethics statement

The PBIDS study protocol was reviewed and approved by the Kenya Medical Research Institute (KEMRI) scientific and ethics review unit (SSC #2761), and the United States Centers for Disease Control and Prevention’s institutional review board (CDC IRB #6775). Written informed consent was obtained from all patients (or parents/guardians if minors), and from compound heads for the household survey.

## Data availability

The data used in this study are available from the KEMRI’s Institutional Data Access / Ethics Committee for researchers who meet the criteria for access to confidential data. The PBIDS data can be accessed by contacting.

## Acknowledgements

We thank all the dedicated staff of KEMRI for their valuable contributions to data and specimen collection, processing and analysis. We are grateful to all study participants, without whom the data would not be accessible. This publication has been approved by the Director of KEMRI. Our work was conducted within the framework of the Research Unit project “Climate Change and Health in Sub-Saharan Africa” funded by the German Research Foundation (DFG/FOR 2936) and the Swiss National Science Foundation under the Weave Lead Agency scheme (SNSF 310030E_186574), with additional funding for disease surveillance studies from the United States Centers for Disease Control and Prevention.

## Disclaimer

The findings and conclusions in this report are those of the authors and do not necessarily represent the official position of United States Centers for Disease Control and Prevention.

## Author information

[1]Swiss Tropical and Public Health Institute, Allschwil, Switzerland; [2]University of Basel, Basel, Switzerland; [3]Kenya Medical Research Institute - Center for Global Health Research, Kisumu, Kenya; [4]United States Centers for Disease Control and Prevention, Nairobi, Kenya; [5]University of Bonn, Bonn, Germany

## Author contributions

Alexis Martin-Makowka and Penelope Vounatsou designed the study and the model experiments. Alexis Martin-Makowka processed and analysed the data, developed the codes and prepared the figures. Alexis Martin-Makowka drafted the manuscript. All authors critically reviewed the manuscript for its intellectual content and approved the final version.

## Appendix 1

### Comprehensive methods

#### Wavelet overview

Wavelets are commonly used for two main purposes. Continuous wavelet transforms allow us to explore the seasonality present in a time series, through descriptive graphs and statistical tests. On the other hand, discrete wavelet transforms algorithmically decompose the time series into subseries of different time scales.

The overall concept of wavelets is related to Fourier analysis and the search for frequencies of a time series (or, inversely, periodicities and seasonalities). However, by its inherent definition, Fourier analysis requires the strong assumption that the time series must be stationary (Daubechies, 1990). In the present study, both the epidemiological data and the climatic data violate this assumption (Pascual et al., 2008), and an alternative approach is therefore needed. Wavelet analysis is a good candidate for this exact reason; unlike the Fourier transform, the wavelet transform does not make use of sine waves to assess the periodicity, but of a ‘wavelet’ instead. As its name suggests, a wavelet is a small, localized in time oscillation. As a result, wavelet analysis is not altered by a change in the seasonal patterns over time, and non-stationary time series can be handled.

By applying a wavelet transform at time-point τ and scale *s* - this latter being related to the notion of Fourier period - to a time series, one could extract information on how strong the seasonal pattern of scale *s* was, around time τ. By proceeding similarly at all time points and scales, we collected valuable information on the seasonal behavior throughout the duration of the study.

#### Continuous wavelet transform

Formally speaking, a wavelet is defined as a function ψ : ℝ → ℂ having at least zero mean and unit energy. Some additional requirements might however be needed for practical purposes (Percival and Walden, 2000). Many wavelets have been developed so far: Daubechies family’s wavelet, Paul’s, Meyer’s, Mexican hat being only a few of them. In our case, we made use of the Morlet wavelet to compute the continuous wavelet transform. The Morlet wavelet has indeed a good trade-off between time and scale resolution (Coulibaly and Burn, 2004; Grinsted et al., 2004). It is complex, and is therefore also suited to the bivariate case, comparing the coherence between two time series (see further)(Rosch and Schmidbauer, 2018). The Morlet Wavelet is defined as follow:

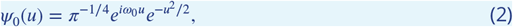

the coefficient ω_0_ was here set to 6 for practical reasons, see e.g. Farge (1992) and Rosch and Schmidbauer (2018).

This equation actually describes the ‘mother’ wavelet; the wavelet of reference. For computing the wavelet transform, we used various stretched and translated versions of the mother wavelet. We obtained the continuous wavelet transform at some time point τ and scale *s* by convolution between the time series and the appropriate wavelet at this time and scale. In other words:

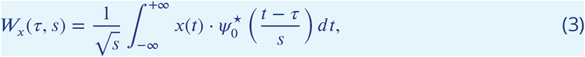

where *W_x_*(τ, *s*) ∈ ℂ is the continuous wavelet transform of the time series *x* at time τ and scale *s*, and the star ^⋆^ defines the complex conjugate.

Based on this quantity, we obtained the wavelet power spectrum at time τ and scale *s*. The formula below was originally from (Torrence and Compo, 1998) but has been updated by (Liu et al., 2007) in order to adjust for scale bias.

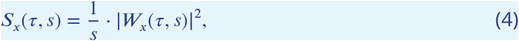

where *S_x_*(τ, *s*) ∈ ℝ represents the wavelet power spectrum and the vertical bars indicates the modulus of a complex number.

The wavelet power spectrum was then plotted using a heat map, sometimes also called a Hovmöller diagram, displaying the power spectrum function of time and scale. The warmer the color, the higher the wavelet power spectrum and so the seasonal pattern. In addition, statistical tests were used to assess the true seasonal behavior. Random lag-1 autoregressive models were simulated to generate a baseline distribution for the wavelet power spectrum. This process allowed us to test the wavelet obtained from our data against the null hypothesis of ‘no seasonal pattern at this time and scale’ (Torrence and Compo, 1998).

Taking the average wavelet power spectrum along a certain period gave the average power spectrum, an unbiased variant of the Fourier transform, which was also tested for seasonality at different period values.

#### Cross-wavelet transform

Based on a similar idea, continuous wavelet transform can be used to plot and test the common patterns in seasonalities between two time series. The so-called cross-wavelet transform is defined by the multiplication of the continuous wavelet transform of both time series as stated by Torrence and Compo (1998), but the formula used here is adjusted with a factor 1/*s* following Veleda et al. (2012).

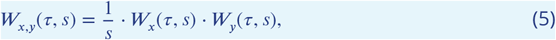

with *W_x_*(τ, *s*), *W_y_*(τ, *s*) the continuous wavelet transform of time series *x, y* as previously defined.

Taking the modulus of this quantity and we obtained the cross-wavelet power spectrum, which could be plotted on a heat map similarly to the wavelet power spectrum:

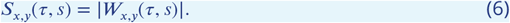

We could statistically assess seasonalities by testing the assumption of no periodicity using two artificially simulated autoregressive lag-1 time series as baseline (Torrence and Compo, 1998). Finally, averaging the cross-wavelet power spectrum returned the average power spectrum, which could also be tested for significance similarly to the average power spectrum.

#### Phase differences

Importantly, the cross-wavelet transform can also assess the phase differences between two time series. The phase difference carries a notion similar to the lag, but not exactly; The underlying information is the time between a peak in the first time series (the leading) and the closest following peak of the other time series (the lagging).

The phase difference angle takes a value *θ* ∈ [0, 2π). This angle represents the proportion of the period between two peaks. As an example, an angle of *θ* = 0 means synchronicity (time series are so-called in-phase), and an angle of *θ* = *π* means the time series are antiphases (see Fig. 1).

Since the definition of phase difference strongly relies on the notion of periodicity, we could only compute reliable phase differences when both time series displayed a periodic/seasonal pattern at this period. Practically, this means that phase differences were only computed for time points and periods where joint seasonality was assessed significant; it was plotted over the cross-wavelet power spectrum heat maps within confidence areas, and took the shape of arrows displaying the angle stated above.

Phase differences could be computed directly from the cross-wavelet transform, as the phase differences have proven to be the argument of the complex number resulting in the computation of the cross-wavelet transform (Rosch and Schmidbauer, 2018):

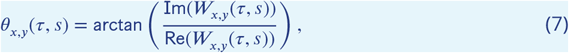

This angle lied within the interval (−π; +π]. It could be converted into a nonnegative angle using the formula:

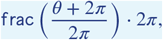

in order to have *θ* ∈ [0; 2π). The function f rac(·) returns the fractional part of the input number.

Arrows plotted in the cross-wavelet plots are displaying the phase difference angles across time and scales. For explicitation, we could use package *WaveletComp* to select a slice of a given period, and plot the phase difference in term of the angle *θ*(*t*) evolving over time. This angle could be converted into month using the formula

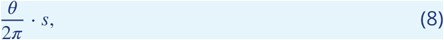

with *θ* the phase difference angle, and *s* the period under investigation. This phase difference could also be turned into a lag, requiring the knowledge of the direction of the relationship between the two variables at this period. A positive relationship would imply that the lag is equal to the phase difference in months. A negative relationship would require to adjust the phase difference by half a period.

#### Maximal overlap discrete wavelet transform

We already had some methods to analyze the time series, for both climate and malaria incidence. However, we did not have a way to state the direction of the relationship between climatic factors and incidence, at a specific period (required to compute the phase differences).

To assess this, we did ‘split’ the time series into different subseries, each having a precise periodicity. Doing so allowed us to use bivariate linear regression over those subseries, to identify more specifically the lag between different time series and get an idea of the relationship between those two.

The following step was then period-dependent subseries decomposition, which could be done using a discrete wavelet transform. As its name may suggest, the discrete wavelet transform is the discretized counterpart of the continuous wavelet transform, even though its application is slightly different. While the continuous wavelet transform carries a lot of information on the seasonality of the time series at any time and scale, the underlying idea of a discrete wavelet transform is to store this information more parsimoniously.

To be more precise, in this situation we made use of the maximal overlap discrete wavelet transform, also sometimes called the decimated wavelet transform, an extension of the classic discrete wavelet transform. Maximal overlap discrete wavelet transform is more redundant, takes slightly more time to compute than discrete wavelet transform, but carries an important feature: a circular shift of the time series induces a circular shift of the maximal overlap discrete wavelet transform, while this statement is not correct for discrete wavelet transform.

A better way to approach the maximal overlap discrete wavelet transform is to see it as a filter. At the lowest scale, we applied this wavelet filter to retain the finest variations in the time series. This subseries, containing only the lower seasonalities, was called *D*_1_, *D* as ‘detail’. We stored it apart and continued with the coarsest scales of the time series, which leaked up through the filter at first stage. We upscaled the wavelet filter so that it retains longer seasonalities in the data. Once again, the filter retained a subseries presenting higher seasonalities, that was called *D*_2_. And we repeated the process again and again until a certain layer *J*. The subseries *D*_1_, *D*_2_, …, *D_J_* were all carrying a particular seasonality. The higher seasonality that could not been caught at this point were carried in a ‘remainder’ subseries called *S_J_*, *S* for ‘smooth’. An interesting property of the discrete wavelet transform is that we can reconstruct the original time series *x*(*t*) out of those subseries as follow:

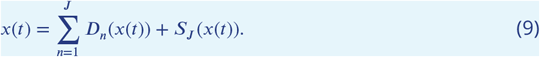

The precise methodology behind this process is quite thorough and falls beyond the scope of this article. A detailed description can however be found in (Percival and Walden, 2000).

Our study made use of the Coiflet wavelet filter of length 30. This wavelet filter had indeed proven efficient over some artificially simulated data of the same nature as the studied data.

After decomposing the time series into multiple subseries, one could investigate the linear relationship between the subseries of climatic and intervention factors and the subseries of malaria incidence. This was done by using a linear regression between the two time series, with a chosen lag comprised between 0 and the maximal meaningful period at this time scale.

## Appendix 2

### Maximal overlap discrete wavelet transforms of climatic time series

**Appendix 2—figure 1.**
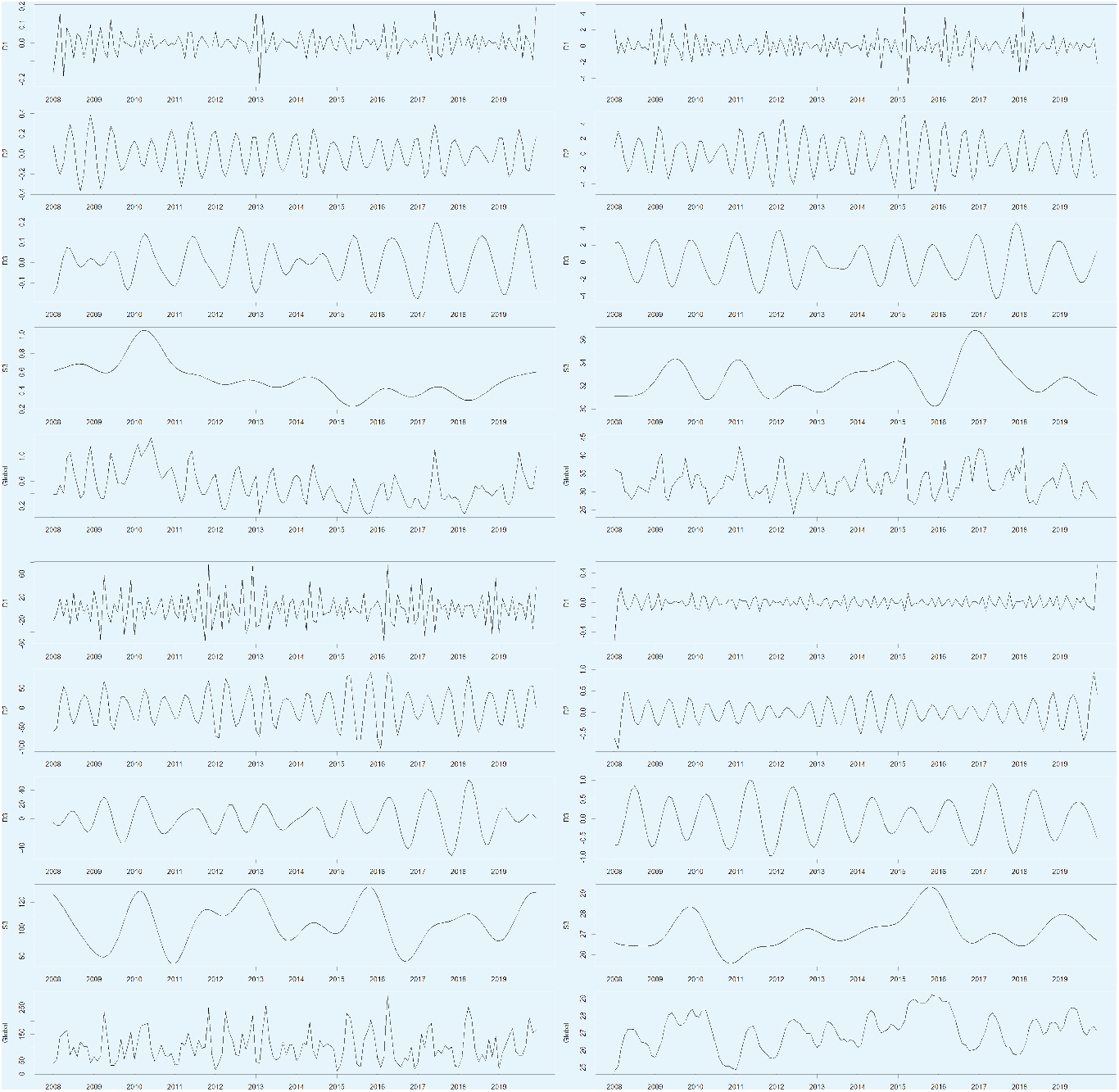
Time series decomposition of the climatic factors, using the maximal overlap discrete wavelet transform. The global (original) time series is plotted at the bottom while the different subseries are plotted above. The subseries *D*_1_ corresponds to time scales of 1-to 4-month, *D*_2_: 4-to 8-month, *D*_3_: 8-to 16-month and *S*_3_ retains seasonalities of periods above 16-month.

## Appendix 3

### Complete table of results for the linear regressions across time scales

**Appendix 3—figure 1.**
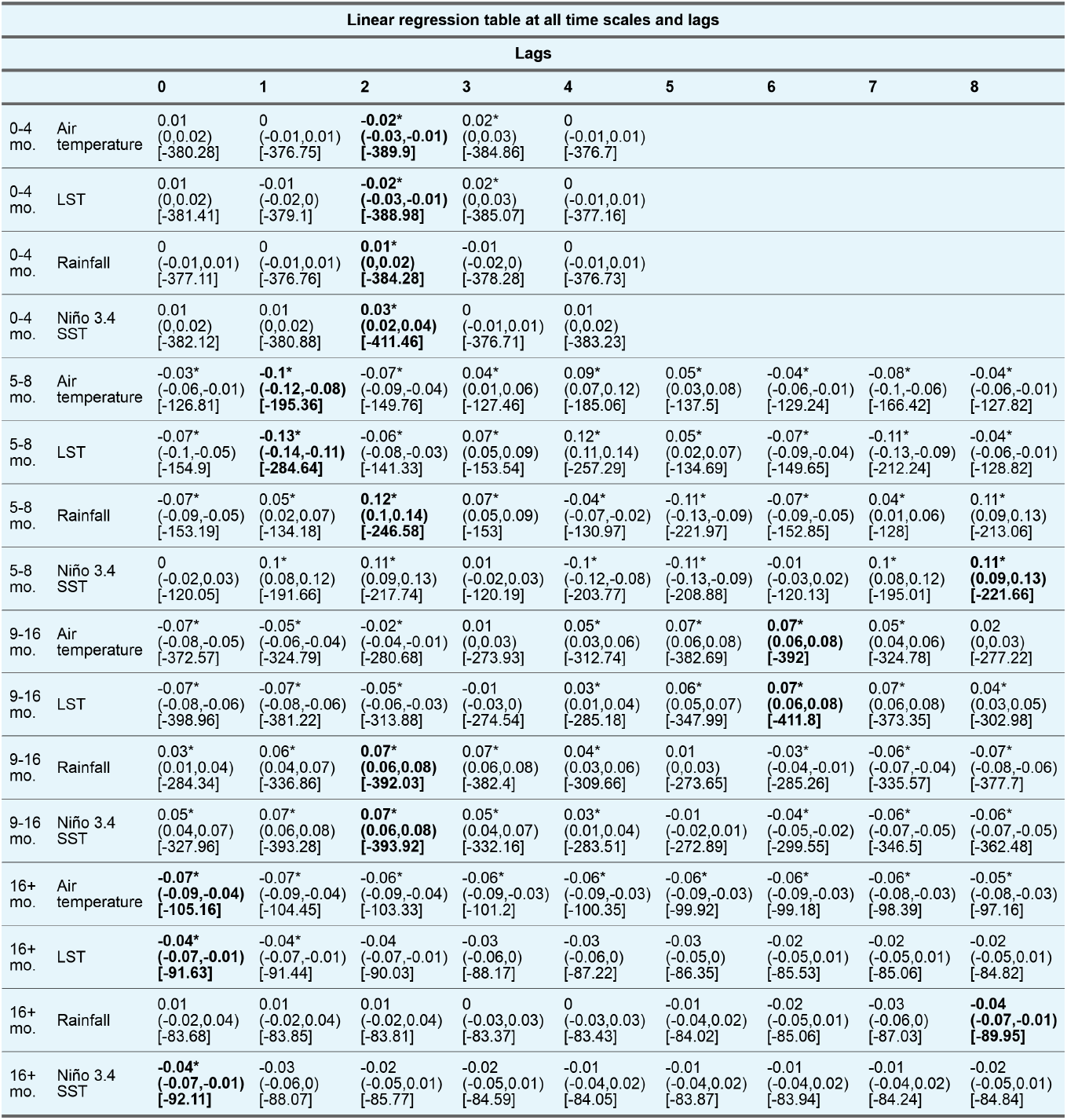
Results of the linear regressions between the malaria incidence time series and the predictors, i.e. the climatic time series at different time scales, for different lags. Maximal overlap discrete wavelet transform-decomposed predictor subseries have been standardized before computation of the model. The first number is the slope coefficient assessing the direction of the relationship. The numbers in brackets represent the 95% confidence interval bounds for the slope coefficient. The last number in square brackets is the Akaike Information Criterion (AIC) of the model. The asterisk means the p-value of the slope coefficient is below the 1% threshold and the bold font highlights the best model for all lags according to minimal AIC. Lags for the lowest time scale (*D*_1_, 0-4-month) have been restricted to a maximum of 4, to avoid lags greater than the time scale.

## References

Anyamba, A., Chretien, J.-P., Britch, S. C., Soebiyanto, R. P., Small, J. L., Jepsen, R., Forshey, B. M., Sanchez, J. L., Smith, R. D., Harris, R., Tucker, C. J., Karesh, W. B., and Linthicum, K. J. (Dec. 2019). “Global disease outbreaks associated with the 2015–2016 El Niño event”. en. In: Scientific Reports 9.1, p. 1930. ISSN: 2045-2322. DOI: 10.1038/s41598-018-38034-z.

Beloconi, A., Nyawanda, B. O., Bigogo, G., Khagayi, S., Obor, D., Danquah, I., Kariuki, S., Munga, S., and Vounatsou, P. (May 2023). “Malaria, climate variability, and interventions: modelling transmission dynamics”. en. In: Scientific Reports 13.1, p. 7367. ISSN: 2045-2322. DOI: 10.1038/s41598-023-33868-8.

Boadu, I. (Jan. 2019). “The changing climate and the changing malaria, the double health challenge”. en. In: International Journal of Advanced Community Medicine 2.1, pp. 05–09. ISSN: 26163586, 26163594. DOI: 10.33545/comed.2019.v2.i1a.02.

Bouma, M. J., Poveda, G., Rojas, W., Chavasse, D., Quinones, M., Cox, J., and Patz, J. (Dec. 1997). “Predicting high-risk years for malaria in Colombia using parameters of El Nino Southern Oscillation”. en. In: Tropical Medicine and International Health 2.12, pp. 1122–1127. ISSN: 1360-2276, 1365-3156. DOI: 10.1046/j.1365-3156.1997.d01-210.x.

Cazelles, B., Cazelles, K., Tian, H., Chavez, M., and Pascual, M. (Sept. 2023). “Disentangling local and global climate drivers in the population dynamics of mosquito-borne infections”. en. In: Science Advances 9.39, eadf7202. ISSN: 2375-2548. DOI: 10.1126/sciadv.adf7202.

Cazelles, B., Champagne, C., and Dureau, J. (Aug. 2018). “Accounting for non-stationarity in epidemiology by embedding time-varying parameters in stochastic models”. en. In: PLoS Computational Biology 14.8. Ed. by K. Koelle, e1006211. ISSN: 1553-7358. DOI: 10.1371/journal.pcbi.1006211.

Cazelles, B., Chavez, M., Magny, G. C. d., Guégan, J.-F., and Hales, S. (Aug. 2007). “Time-dependent spectral analysis of epidemiological time-series with wavelets”. en. In: Journal of the Royal Society Interface 4.15, pp. 625– 636. ISSN: 1742-5689, 1742-5662. DOI: 10.1098/rsif.2007.0212.

Coulibaly, P. and Burn, D. H. (Mar. 2004). “Wavelet analysis of variability in annual Canadian streamflows: Wavelet Analysis of Variability”. en. In: Water Resources Research 40.3. ISSN: 00431397. DOI: 10.1029/2003WR002667.

Daubechies, I. (Sept. 1990). “The wavelet transform, time-frequency localization and signal analysis”. en. In: IEEE Transactions on Information Theory 36.5, pp. 961–1005. ISSN: 00189448. DOI: 10.1109/18.57199.

Farge, M. (1992). “Wavelet transforms and their applications to turbulence”. en. In: Annu. Rev. Fluid Mech. 24, pp. 395–457.

Field, C. B. and Barros, V. R. (2014). Climate change 2014: impacts, adaptation, and vulnerability Working Group II contribution to the fifth assessment report of the Intergovernmental Panel on Climate Change. en. Tech. rep. New York.

Funk, C., Peterson, P., Landsfeld, M., Pedreros, D., Verdin, J., Shukla, S., Husak, G., Rowland, J., Harrison, L., Hoell, A., and Michaelsen, J. (Dec. 2015). “The climate hazards infrared precipitation with stations—a new environmental record for monitoring extremes”. en. In: Scientific Data 2.1, p. 150066. ISSN: 2052-4463. DOI: 10.1038/sdata.2015.66.

Grinsted, A., Moore, J. C., and Jevrejeva, S. (Nov. 2004). “Application of the cross wavelet transform and wavelet coherence to geophysical time series”. en. In: Nonlinear Processes in Geophysics 11.5/6, pp. 561–566. ISSN: 1607-7946. DOI: 10.5194/npg-11-561-2004.

Kapwata, T., Wright, C. Y., Preez, D.J. du, Kunene, Z., Mathee, A., Ikeda, T., Landman, W., Maharaj, R., Sweijd, N., Minakawa, N., and Blesic, S. (Oct. 2021). “Exploring rural hospital admissions for diarrhoeal disease, malaria, pneumonia, and asthma in relation to temperature, rainfall and air pollution using wavelet transform analysis”. en. In: Science of The Total Environment 791, p. 148307. ISSN: 00489697. DOI: 10.1016/j.scitotenv.2021.148307.

Lafferty, K. D. (Apr. 2009). “The ecology of climate change and infectious diseases”. en. In: Ecology 90.4, pp. 888– 900. ISSN: 0012-9658. DOI: 10.1890/08-0079.1.

Liu, Y., San Liang, X., and Weisberg, R. H. (Dec. 2007). “Rectiflcation of the bias in the wavelet power spectrum”. en. In: Journal of Atmospheric and Oceanic Technology 24.12, pp. 2093–2102. ISSN: 1520-0426, 0739-0572. DOI: 10.1175/2007JTECHO511.1.

Mordecai, E. A., Caldwell, J. M., Grossman, M. K., Lippi, C. A., Johnson, L. R., Neira, M., Rohr, J. R., Ryan, S. J., Savage, V., Shocket, M. S., Sippy, R., Ibarra, A. M. S., Thomas, M. B., and Villena, O. (2019). “Thermal biology of mosquito-borne disease”. en. In: Ecology Letters 22.10. _eprint: https://onlinelibrary.wiley.com/doi/pdf/10.1111/ele.13 pp. 1690–1708. ISSN: 1461-0248. DOI: 10.1111/ele.13335.

Muñoz Sabater, J. (2019). ERA5-land hourly data 1981 to present.

Nyawanda, B. O., Beloconi, A., Khagayi, S., Bigogo, G., Obor, D., Otieno, N. A., Lange, S., Franke, J., Sauerborn, R., Utzinger, J., Kariuki, S., Munga, S., and Vounatsou, P. (May 2023). “The relative effect of climate variability on malaria incidence after scale-up of interventions in western Kenya: a time-series analysis of monthly incidence data from 2008 to 2019”. en. In: Parasite Epidemiology and Control 21, e00297. ISSN: 24056731. DOI: 10.1016/j.parepi.2023.e00297.

Ototo, E. N., Githeko, A. K., Wanjala, C. L., and Scott, T. W. (Dec. 2011). “Surveillance of vector populations and malaria transmission during the 2009/10 El Niño event in the western Kenya highlands: opportunities for early detection of malaria hyper-transmission”. en. In: Parasites & Vectors 4.1, p. 144. ISSN: 1756-3305. DOI: 10.1186/1756-3305-4-144.

Pascual, M., Cazelles, B., Bouma, M., Chaves, L., and Koelle, K. (Jan. 2008). “Shifting patterns: malaria dynamics and rainfall variability in an African highland”. en. In: Proceedings of the Royal Society B: Biological Sciences 275.1631, pp. 123–132. ISSN: 0962-8452, 1471-2954. DOI: 10.1098/rspb.2007.1068.

Percival, D. B. and Walden, A. T. (2000). Wavelet methods for time series analysis. en. Cambridge: Cambridge University Press. ISBN: 978-0-511-84104-0. DOI: 10.1017/CBO9780511841040.

Rayner, N. A., Parker, D. E., Horton, E. B., Folland, C. K., Alexander, L. V., Rowell, D. P., Kent, E. C., and Kaplan, A. (2003). “Global analyses of sea surface temperature, sea ice, and night marine air temperature since the late nineteenth century”. en. In: Journal of Geophysical Research 108.D14, p. 4407. ISSN: 0148-0227. DOI: 10.1029/2002JD002670.

Reiner, R. C., Geary, M., Atkinson, P. M., Smith, D. L., and Gething, P. W. (Dec. 2015). “Seasonality of Plasmodium falciparum transmission: a systematic review”. en. In: Malaria Journal 14.1, p. 343. ISSN: 1475-2875. DOI: 10.1186/s12936-015-0849-2.

Rosch, A. and Schmidbauer, H. (2018). WaveletComp 1.1: A guided tour through the R package. en.

Tanser, F. C., Sharp, B., and Sueur, D. le (Nov. 2003). “Potential effect of climate change on malaria transmission in Africa”. en. In: Lancet 362.9398, pp. 1792–1798. ISSN: 01406736. DOI: 10.1016/S0140-6736(03)14898-2.

Thomson, M. C., Ukawuba, I., Hershey, C. L., Bennett, A., Ceccato, P., Lyon, B., and Dinku, T. (Sept. 2017). “Using rainfall and temperature data in the evaluation of national malaria control programs in Africa”. en. In: American Journal of Tropical Medicine and Hygiene 97.3_Suppl, pp. 32–45. ISSN: 0002-9637, 1476-1645. DOI: 10.4269/ajtmh.16-0696.

Torrence, C. and Compo, G. P. (1998). “A practical guide to wavelet analysis”. en. In: Bulletin of the American Meteorological Society 79.1, p. 18.

Trenberth, K. E. (Dec. 1997). “The Definition of El Niño”. en. In: Bulletin of the American Meteorological Society 78.12, pp. 2771–2777. ISSN: 0003-0007, 1520-0477. DOI: 10.1175/1520-0477(1997)078<2771:TDOENO>2.0.CO;2.

Veleda, D., Montagne, R., and Araujo, M. (Sept. 2012). “Cross-wavelet bias corrected by normalizing scales”. en. In: Journal of Atmospheric and Oceanic Technology 29.9, pp. 1401–1408. ISSN: 0739-0572, 1520-0426. DOI: 10.1175/JTECH-D-11-00140.1.

Wan, Z., Hook, S., and Hulley, G. (2015). “MOD11A2 MODIS/Terra land surface temperature/emissivity 8-day L3 global 1km SIN grid V006”. en. In: Nasa Eosdis Land Processes Daac. NASA LP DAAC 10.10.5067.

Wang, Z., Liu, Y., Li, Y., Wang, G., Lourenço, J., Kraemer, M., He, Q., Cazelles, B., Li, Y., Wang, R., Gao, D., Li, Y., Song, W., Sun, D., Dong, L., Pybus, O. G., Stenseth, N. C., and Tian, H. (Apr. 2022). “The relationship between rising temperatures and malaria incidence in Hainan, China, from 1984 to 2010: a longitudinal cohort study”. en. In: Lancet Planetary Health 6.4, e350–e358. ISSN: 25425196. DOI: 10.1016/S2542-5196(22)00039-0.

WHO (2023). World malaria report 2023. en. Tech. rep. Geneva: World Health Organization.

